# Calculating epidemiological outcomes from simulated longitudinal data

**DOI:** 10.1101/2025.04.30.25326766

**Authors:** Selina Pi, Jeremy D. Goldhaber-Fiebert, Fernando Alarid-Escudero

## Abstract

Microsimulation models generate individual life trajectories that must be summarized as population-level outcomes for model calibration and validation. While there are established formulas to calculate outcomes such as prevalence, incidence, and lifetime risk from cross-sectional and short-term longitudinal studies, limited guidance exists to calculate these outcomes using long-term longitudinal data due to the rarity of large-scale studies covering events across the human lifespan. This technical report presents various methods to calculate epidemiological outcomes from simulated longitudinal data, from replicating a real-world study design to fully incorporating longitudinal disease and exposure durations. We provide an open-source code base with functions in R to calculate the prevalence, incidence, age-conditional risk, lifetime risk, and disease-specific mortality of a condition from individual-level time-to-event data. In addition, we provide guidance and code for calculating cancer-related outcomes from individual-level data, such as the stage distribution at diagnosis, the distribution of precancerous lesion multiplicity, and the mean dwell and sojourn time. Given the various possible formulations for certain outcomes, we call for increased transparency in reporting how summary outcomes are derived from microsimulation model outputs, and we anticipate that this report will facilitate the calculation of epidemiological outcomes in both simulated and real-world data.

## 1 Introduction

Calibration and validation of a disease natural history model center around ensuring that the model outputs match relevant epidemiological endpoints, such as incidence, prevalence, lifetime risk, and disease-specific mortality [1]. These statistics may be derived from screening studies or disease surveillance data. While formulas for prevalence and incidence have been described for population-level differential equation models [2, 3, 4, 5, 6], there is limited guidance on calculating such summary outcomes from simulated time-to-event data, possibly because longitudinal data analogous to the life trajectories generated by an individual-level simulation model are rare in practice. Prevalence and incidence have been used as calibration targets for several microsimulation models (e.g., [7, 8, 9, 10, 11, 12, 13]), but their documentation does not specify how these outcomes were calculated from the model outputs. This technical report provides mathematical formulas extrapolating cross-sectional and short-term longitudinal outcome definitions to the lifetime setting for simulated individual trajectories. We demonstrate that there are multiple ways to calculate certain outcomes from lifetime data, with tradeoffs in computational expense, precision, and bias. Therefore, we argue that it is important for researchers to transparently report how these outcomes are calculated for microsimulation models. In addition to prevalence and incidence, we discuss how to calculate age-conditional and lifetime risk, disease-specific mortality, and cancer-related outcomes, such as the stage distribution at diagnosis, the distribution of the number of lesions or tumors, and the mean dwell or sojourn time. Functions to calculate outcomes for one-time conditions from a matrix of life trajectories are provided in an open-source repository (https://github.com/sjpi22/microsumulation/).

## 2 Outcomes

We define variables for event times and other notation used across this report in Table 1. Event episodes for recurrent conditions are assumed not to overlap, so 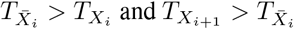. We assume that there are no missing values. For events that would have occurred after an individual’s death or events for which a person is not at risk, the event time is assumed to be infinite. Censoring can be due to death, loss to follow-up, or loss of eligibility, for example due to the diagnosis of a condition prior to a screening study of asymptomatic individuals. For the purpose of calibration, when simulating individual life histories, care should be taken to ensure that the simulated population is representative (e.g., has the same baseline characteristics) as that of the empirical studies from which the calibration target of interest is derived. The formulas assume a cohort of *N* life histories representative of the population or subgroups for which outcomes will be calculated.

**Table 1:**
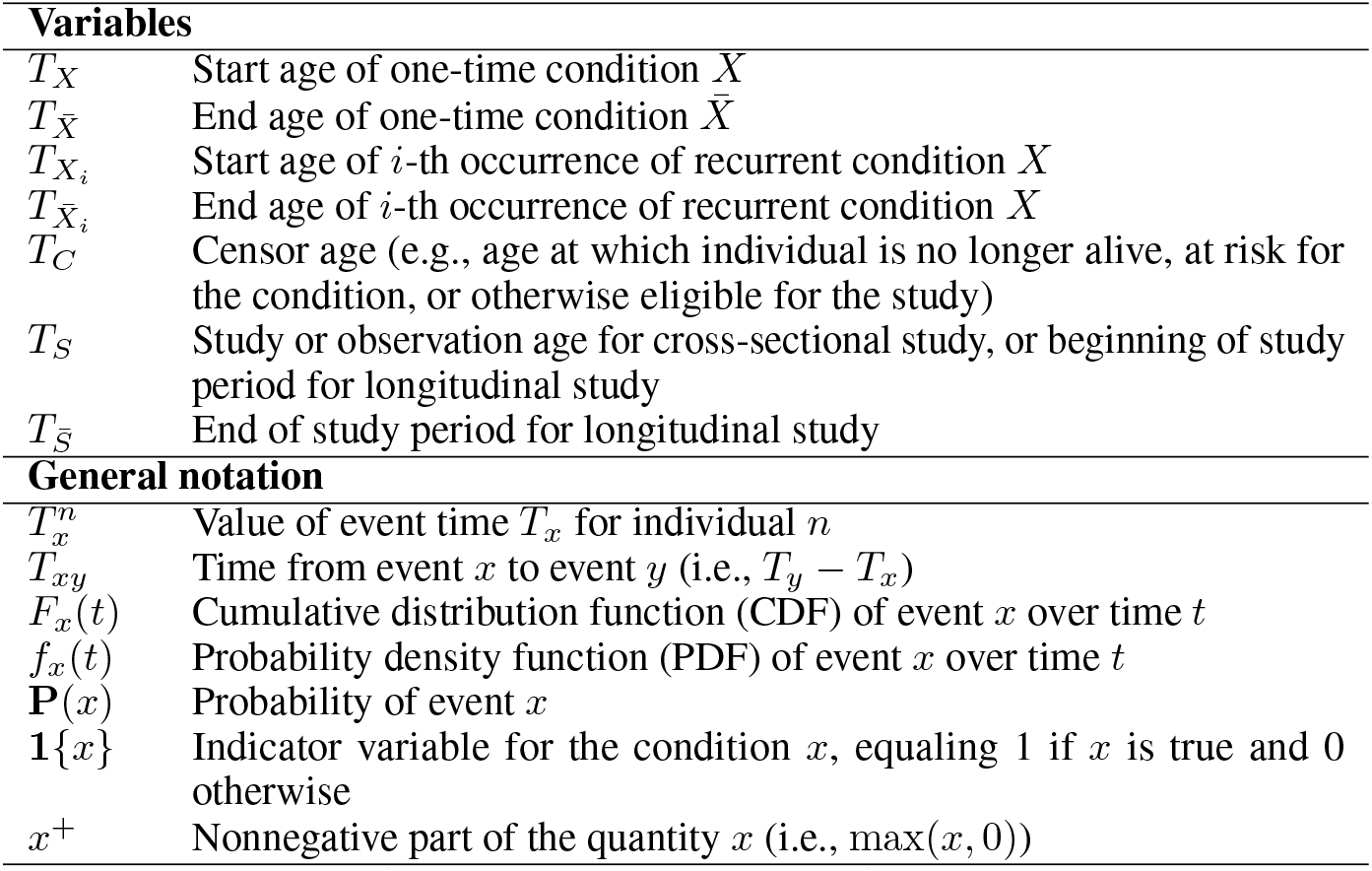
Notation for events and quantities.

| **Variables** |  |
| --- | --- |
| $T_X$ | Start age of one-time condition $X$ |
| $T_{\bar{X}}$ | End age of one-time condition $\bar{X}$ |
| $T_{X_i}$ | Start age of $i$-th occurrence of recurrent condition $X$ |
| $T_{\bar{X}_i}$ | End age of $i$-th occurrence of recurrent condition $X$ |
| $T_C$ | Censor age (e.g., age at which individual is no longer alive, at risk for the condition, or otherwise eligible for the study) |
| $T_S$ | Study or observation age for cross-sectional study, or beginning of study period for longitudinal study |
| $T_{\bar{S}}$ | End of study period for longitudinal study |
| **General notation** |  |
| $T_x^n$ | Value of event time $T_x$ for individual $n$ |
| $T_{xy}$ | Time from event $x$ to event $y$ (i.e., $T_y - T_x$) |
| $F_x(t)$ | Cumulative distribution function (CDF) of event $x$ over time $t$ |
| $f_x(t)$ | Probability density function (PDF) of event $x$ over time $t$ |
| $\mathbf{P}(x)$ | Probability of event $x$ |
| $\mathbf{1}\{x\}$ | Indicator variable for the condition $x$, equaling 1 if $x$ is true and 0 otherwise |
| $x^+$ | Nonnegative part of the quantity $x$ (i.e., $\max(x, 0)$) |

### 2.1 Prevalence

The prevalence of a condition is defined as the proportion of individuals with the condition out of the population at risk [14]. In cross-sectional studies and disease surveillance reporting, prevalence is often stratified by age groups to reflect age-specific differences in risk, though practical and ethical considerations often preclude reporting exact individual ages [15]. In terms of probabilities P(*x*), the true prevalence *P* (*t*) of a one-time condition *X* at *t* according to the notation in Table 1 is:

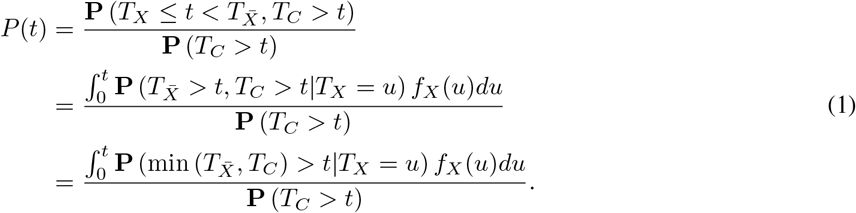

When censoring is independent of the condition and the condition does not end before the censor time, Equation 1 simplifies to *F*_*X*_ (*t*). Assuming no birth cohort effects, the true prevalence *P* (*t*_1_, *t*_2_) of the condition from age *t*_1_ to *t*_2_ is the expected value of the single-age prevalence from *t*_1_ to *t*_2_, weighted by the proportion of the population that is uncensored:

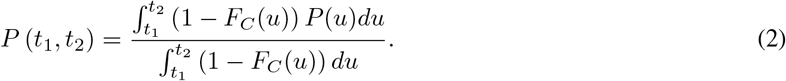

In this section, we provide three approaches to calculate age-specific disease prevalence from simulated life trajectories and compare the Monte Carlo variation, computation time, and bias from the true prevalence associated with each method.

#### 2.1.1 Cross-sectional formulation (point prevalence)

Age-specific prevalence can be calculated for a simulated cohort in a cross-sectional manner analogous to real-world studies by sampling a study or observation age *T*_*S*_ for each individual, excluding individuals who are censored before *T*_*S*_ or otherwise ineligible for the study, and calculating the proportion of at-risk individuals in the age range of interest with the condition at *T*_*S*_. *T*_*S*_ can be determined by assigning birth years *Y*_*birth*_ representative of the population, setting a study year or sampling from a range of years *Y*_*S*_, and calculating *T*_*S*_ = *Y*_*S*_ − *Y*_*birth*_. Alternately, *T*_*S*_ can be randomly sampled between the age ranges of interest. A uniform distribution between the minimum and maximum ages for which prevalence will be calculated would provide a representative estimate of the population in terms of the relative frequency of individuals at each age who have not been censored. To calculate the cross-sectional or point prevalence *P*_*cs*_ of a condition *X* between age *t*_1_ (inclusive) to *t*_2_ (exclusive), we use the following formula across the simulated life trajectories *n* and event episodes *i*:

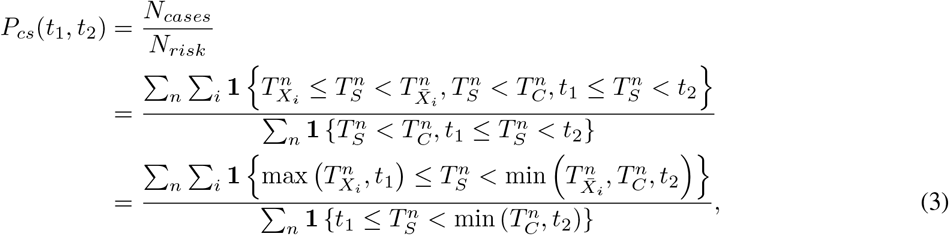

where *N*_*cases*_ indicates the number of individuals with the condition at *T*_*S*_ among the *N*_*risk*_ at-risk individuals observed between age *t*_1_ and *t*_2_. For a one-time condition, Equation 3 simplifies to

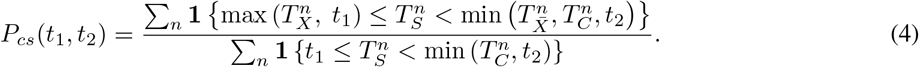

As an example, for the prevalence of colorectal cancer (CRC) in a screening study (e.g., among those without diagnosed CRC) of average-risk individuals, *T*_*X*_ would be the age at CRC onset, 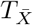 would be the age at the symptomatic detection of CRC, and *T*_*C*_ would be the earliest of the age of death and the age of symptomatic CRC diagnosis in Equation 4. We assume that individuals with inflammatory bowel disease, familial adenomatous polyposis, and other conditions associated with an increased risk of CRC are already excluded. In this case, 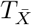 is redundant to *T*_*C*_, but this does not always occur; for instance, for the prevalence of precancerous lesions without CRC in the same study, *T*_*X*_ would be the age at which the first lesion develops,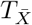 would be the age at onset of CRC (assuming no lesion regression), and *T*_*C*_ would be the same as for preclinical CRC prevalence.

To calculate the prevalence of a condition at a single age *t, T*_*S*_, *t*_1_, and *t*_2_ are all set to *t*, and prevalence is calculated as the number of individuals with the condition out of the at-risk individuals at age *t*, making Equation 3 simplify to:

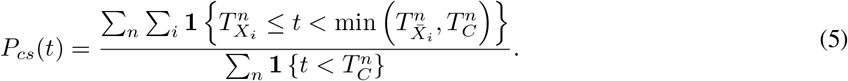

Conceptualizing prevalence as a proportion, the Wilson score interval with continuity correction can be used to calculate confidence intervals (CIs) for cross-sectional prevalence [16]. To estimate the standard error (SE), we can use the binomial approximation:

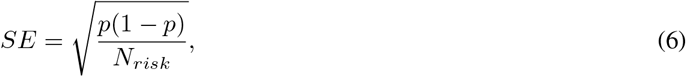

where *p* is the prevalence estimate and *N*_*risk*_ is the number of individuals in the denominator for the estimate.

#### 2.1.2 Longitudinal formulation

Assuming negligible birth cohort effects within the age intervals of interest, we can make more efficient use of the simulated longitudinal data by calculating the prevalence *P*_*long*_ of a condition *X* from age *t*_1_ to *t*_2_ as the proportion of time at risk in the age range that individuals have the condition:

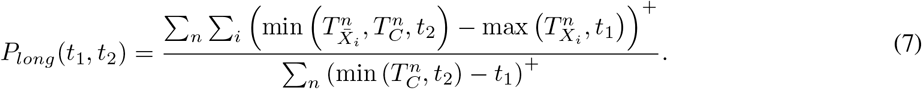

#### 2.1.3 Repeated cross-sectional formulation

The repeated cross-sectional (RCS) formulation of prevalence is a hybrid of the cross-sectional and longitudinal approaches that sums the numerators and denominators of the single-age prevalence (Equation 5) at each integer age from *t*_1_ to *t*_2_ and is well-suited to discrete-time microsimulation outputs:

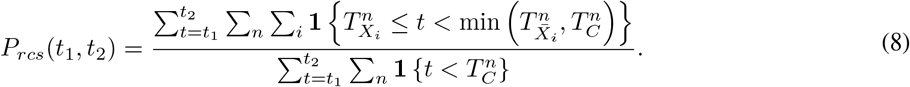

#### 2.1.4 Comparison of formulations

We compare the three formulations with a simulation study in which half of the population is at risk of developing a condition *X* whose onset time follows a Weibull distribution with shape 4 and scale 60. Individuals die uniformly between age 30 and 100 independently of the condition, and the condition lasts until death. Therefore, the true prevalence of the condition at any time point is merely half value of the Weibull CDF at that point (see Figure 1), and the true prevalence in an age range is calculated using Equation 2 with *P* (*u*) = *F*_*X*_ (*u*). We simulate 1,000 cohorts of size 100,000 and calculate the cross-sectional, longitudinal, and repeated cross-sectional prevalence of the condition across the population from age 30 to 80 and within 10-year age ranges from 30 to 80. The top two panels of Figure 2 show the mean estimate for each formulation in terms of the percentage difference from the true prevalence (e.g., (estimated *−* true)*/*true), and the error bars represent the standard deviation of the 1,000 estimates as a proportion of the true prevalence. The bottom two panels reflect the mean time per calculation.

**Figure 1:**
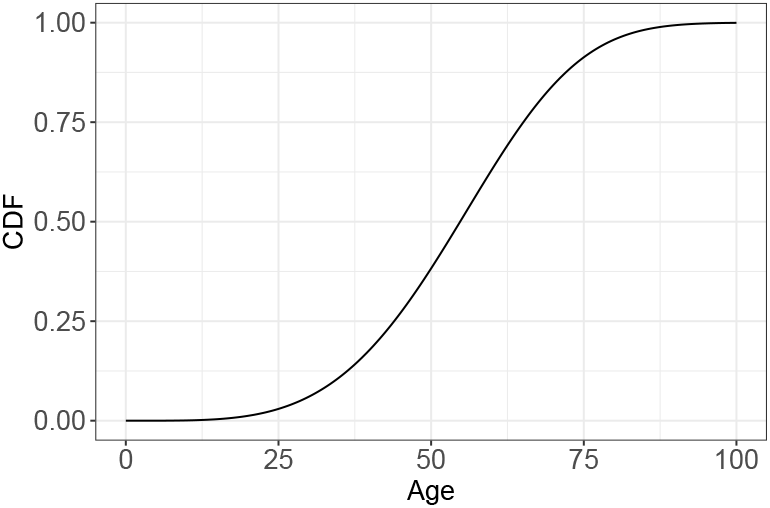
Cumulative distribution function for Weibull distribution with shape 4 and scale 60

**Figure 2:**
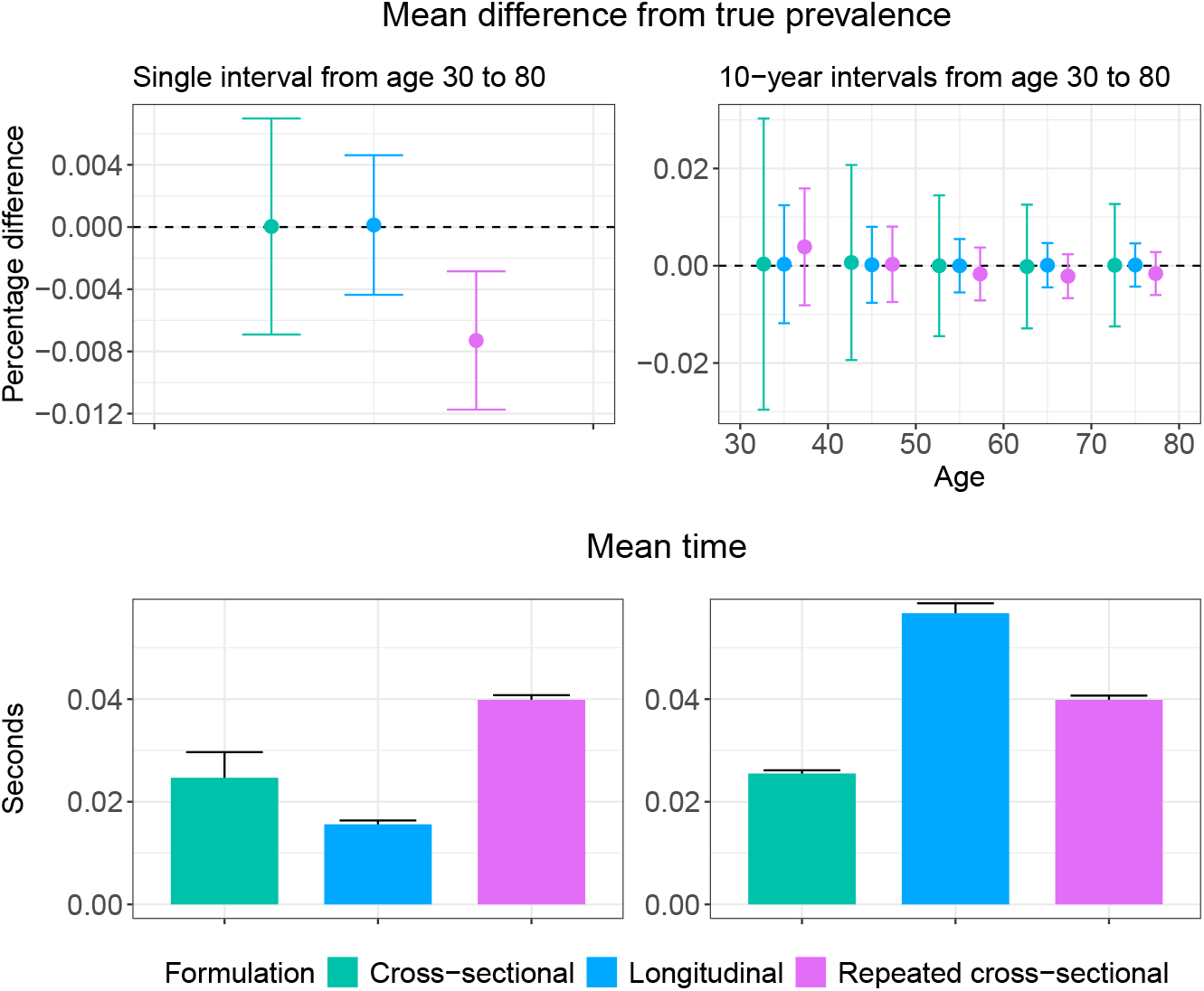
Comparison of formulations for aggregate prevalence from age 30 to 80 (left) and prevalence in 10-year intervals from age 30 to 80 (right), with standard deviation of the estimates shown as error bars

To calculate prevalence in a single age range, the longitudinal formulation is fastest with the lowest Monte Carlo variation. The repeated cross-sectional method demonstrates considerable bias for a large age range, as the true prevalence is not within one standard deviation of the mean estimate. For a typical use case, in which prevalence is calculated for several smaller age ranges, the longitudinal formulation exceeds the other two methods in computation time but produces less than half the Monte Carlo variation of the cross-sectional method. The repeated cross-sectional method is faster and has similar variance as the longitudinal method. It is on average slightly biased from the true prevalence, though the magnitude of bias is lower than the standard deviation of the estimates.

Overall, the longitudinal method has the optimal level of Monte Carlo variation and bias, but its computation time increases linearly with the number of age brackets, whereas the computation times of the cross-sectional and repeated cross-sectional methods appear to be stable with the number of age ranges. The repeated cross-sectional method has more precision than the cross-sectional method and is faster than the longitudinal method when there is a sufficient number of age brackets, but it can produce biased estimates when the single-age prevalence function has a high degree of curvature. Given how it is calculated, we hypothesize that the computation time increases linearly with the width of the full age range to calculate prevalence. To choose which formulation to use, we recommend assessing the tradeoffs between the computation time, necessary precision, and amount of bias across the three methods for the specific use case and have provided template code in the repository to reproduce Figure 2 to do so.

### 2.2 Incidence

Incidence is defined as the number of events per person-year at risk [17]. For age-specific incidence, the denominator is the total person-years at risk within the age range of interest [18]. The time required to observe new cases means that incidence cannot be measured in cross-sectional studies [19]. The Surveillance, Epidemiology, and End Results (SEER) Program calculates the crude incidence rate as the number of new cases or events in a given year divided by the total population, or the total population of a given sex for sex-specific cancers [20]. Cancer recurrences are generally not included as incident cases [21]. For rare conditions with a low incidence, the rate is often reported per 100,000. Here, we show formulas for crude incidence rates, which are acceptable for the narrow age ranges for which cancer incidence targets are typically reported (e.g., 5- or 10-year age ranges). For the modifications needed to obtain age-adjusted incidence rates across a full population, we refer to [20].

With lifetime data, we can calculate the incidence of a condition between age *t*_1_ and *t*_2_ from simulated life trajectories *n* and event episodes *i* as:

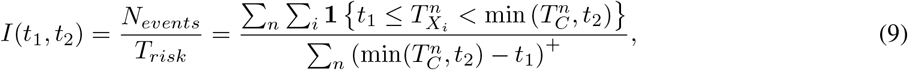

where *N*_*events*_ is the number of new condition onset events from *t*_1_ to *t*_2_ and *T*_*risk*_ is the total person-years at risk from *t*_1_ to *t*_2_. For a one-time condition, Equation 9 simplifies to:

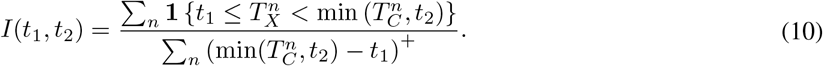

As an example, to calculate the incidence rate of cancer diagnoses among individuals not previously diagnosed, *T*_*X*_ would be the age at cancer diagnosis and *T*_*C*_ would be the earliest of the ages at death and cancer diagnosis in Equation 10. For the incidence of cancer diagnoses out of the total population, *T*_*C*_ would be the age of death.

To create an analog of a real-world longitudinal study, each individual would have a study or observation period from *T*_*S*_ to 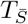, and Equation 9 would be modified by bounding the event and exposure times by the observation period, as described in [18]:

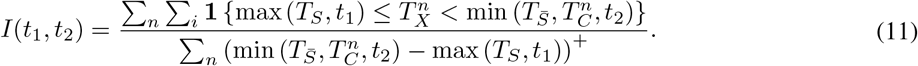

Following [20], assuming that the annual rate of cases is Poisson-distributed, the SE of the incidence rate of a one-time condition is calculated as:

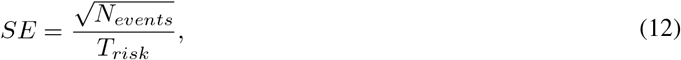

and the bounds of the (1 − *α*) × 100% CIs are calculated as:

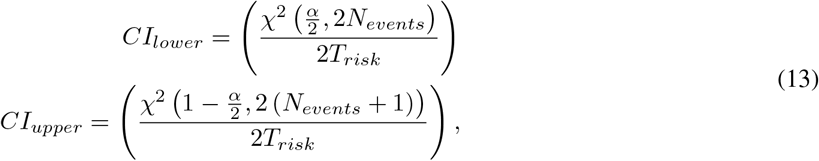

where *N*_*events*_ and *T*_*risk*_ are as defined for Equation 9, and *χ*^2^ (*α, ν*) is the value *x* at which the chi-squared distribution with *ν* degrees of freedom has a probability *α* of being greater than *x*.

### 2.3 Age-conditional and lifetime risk

The risk of developing a condition *X* for the first time between age *t*_1_ and *t*_2_ conditional on not having developed the condition before *t*_1_ is calculated as:

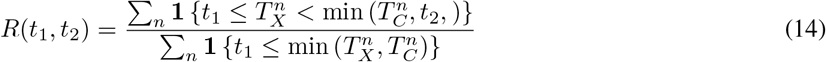

The lifetime risk of the condition can be calculated by setting *t*_1_ = 0 and *t*_2_ = *∞*, making Equation 14 simplify to:

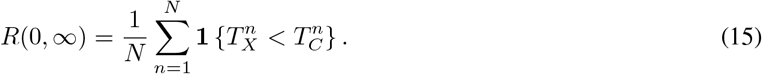

### 2.4 Disease-specific mortality

For a microsimulation model in which the cause of death is simulated, we can use standard competing risk methods to calculate the disease-specific mortality and survival rates from diagnosis. We fit a Kaplan-Meier curve with event time *T*_*XC*_ = *T*_*C*_ − *T*_*X*_ and event indicator *D* to the population for which *T*_*X*_ ≤ *T*_*C*_, where *T*_*X*_ is the time at condition onset, *T*_*C*_ is the time of death, and *D* is 1 if a simulated individual’s death is due to the condition and 0 if the death is due to other causes. The population can first be filtered to subgroups such as individuals diagnosed at a particular age or stage of disease. The Kaplan-Meier curve provides the proportion that have not died of cancer *t* years after diagnosis.

### 2.5 Stage distribution of cancer

The stage distribution of cancer is the proportion diagnosed at each stage among individuals diagnosed with cancer in their lifetimes. Letting *T*_*X*_ be the time at the diagnosis of cancer, *T*_*C*_ be the time of death, and *S*(*T*_*X*_) be the stage at diagnosis, the proportion of individuals with cancer diagnosed at stage *s* is calculated as:

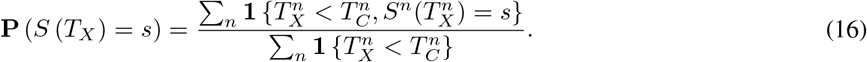

### 2.6 Lesion multiplicity

The distribution of precancerous lesion multiplicity is the proportion of individuals with a given number of concurrent lesions among individuals with at least one lesion. Letting *L*(*t*_1_, *t*_2_) be the number of concurrent lesions at an observation time between *t*_1_ to *t*_2_, 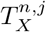 and 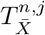 respectively be the times at which lesion *j* in individual *n* develops and ends, 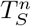 be the age at which an individual is observed for lesions (e.g., through screening), and 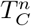 be the earliest of the time of death or the time at which cancer first develops from any lesion for individual *n*, we calculate the proportion of individuals with *x* lesions among those with any lesions as:

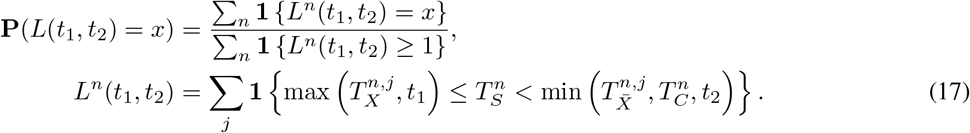

The distribution can also be calculated longitudinally by converting the lesion-level duration data to mutually exclusive episodes with a single number of lesions. Similar calculations can be used for other characteristics of precancerous lesions and tumors, such as the distribution of the largest lesion size.

### 2.7 Dwell and sojourn time

Modeling may help estimate intermediate outcomes of cancer natural history that are difficult to observe. These outcomes include sojourn time, defined as the time from the onset to the diagnosis of cancer; precancerous lesion dwell time, or the time from precancerous lesion onset to the onset of preclinical cancer; and stage-specific dwell time, or the time from one stage of cancer to the next. The mean duration generally follows the formula:

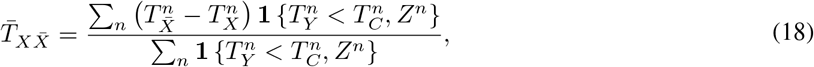

where *T*_*Y*_ is an event that must occur before the censor time for an individual to be included in the statistic and *Z* is a catch-all for any other inclusion criteria. For the mean sojourn time among individuals diagnosed with cancer in their lifetimes, *T*_*X*_ is the onset time of preclinical cancer, 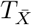 is the time at diagnosis of cancer, *T*_*Y*_ is also the time at diagnosis, *T*_*C*_ is the time of death, and *Z* contains no restrictions. For cancers that include a precursor lesion state, the mean dwell time of lesions among individuals diagnosed with cancer is calculated with *T*_*X*_ equal to the onset time of the first lesion,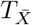 equal to the onset of preclinical cancer assuming no lesion regression, and *T*_*Y*_, *T*_*C*_, and *Z* equal to the same quantities as for mean sojourn time. For the dwell time from stage *s* to *s* + 1 of cancer before diagnosis among individuals diagnosed at or after stage *s* + 1, *T*_*X*_ equals the start time of stage *s*, 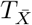 equals the start time of stage *s* + 1, *T*_*Y*_ is the time at diagnosis, *T*_*C*_ is the time of death, and *Z* restricts to individuals diagnosed at stage *s* + 1 or later.

## 3 Discussion

In this technical report, we show how to map longitudinal event data as produced by individual-level models to population outcomes that can be compared with statistics typically reported in real-world epidemiological studies, such as prevalence, incidence, and age-conditional risk. In addition, we provide equations for other useful statistics from microsimulation model outputs, such as the stage distribution of cancer and the mean duration of a health state. The formulas for these summary statistics will also facilitate likelihood computation for model calibration. Prevalence can be calculated either by replicating a cross-sectional study or using longitudinal condition and risk durations as inputs, which results in lower Monte Carlo variation but may be more computationally expensive. The precision gains of the longitudinal formulation compared to the cross-sectional formulation essentially come from a longer observation period and from allowing individuals to contribute data to multiple age groups. When calculating analogs of real-world targets with simulated longitudinal data, care should be taken to define event time inputs and other eligibility criteria for the simulated population consistent with the comparator study. Given the multiple possible formulations and the sensitivity of outcomes to the characteristics of the study population, we recommend that modelers report equations, functions, or both for epidemiological outcome calculations used for microsimulation models.

## Data Availability

All data produced are available online at

https://github.com/sjpi22/microsumulation/tree/main

## 4 Acknowledgments

SP was supported by the US National Institutes of Health (NIH Grant T15LM007033) and the National Science Foundation (NSF) Graduate Research Fellowship Program (Grant DGE-2146755). FAE was supported by grants U01-CA253913 and U01-CA265750 from the National Cancer Institute (NCI) as part of the Cancer Intervention and Surveillance Modeling Network (CISNET). Any opinions, findings, and conclusions or recommendations expressed in this material are those of the authors and do not necessarily reflect the views of the NIH, NSF, or NCI. Authors have no competing interests to declare.

